# Optimizing Ambulatory Groin Hernia Repair in Public Healthcare Frameworks: A Prospective Analysis of Predictive Factors for Discharge Failure

**DOI:** 10.64898/2026.05.27.26354207

**Authors:** Jihen Krichen, Asma Sghaier, Rana Dhouib, Slah Souii, Maissa Tioumi, Sihem Sindi, Bilel Faidi, Khalil Ben Salah

**Affiliations:** Department of Visceral and Digestive Surgery, Ibn Al Jazzar University Hospital, Aghlabids’ Unit, Kairouan, Tunisia; Department of Visceral and Digestive Surgery, Farhat Hached University Hospital, Sousse, Tunisia; Department of Anesthesia and Surgical Intensive Care, Ibn Al Jazzar University Hospital, Aghlabids’ Unit, Kairouan, Tunisia; Faculty of Medicine of Sousse, University of Sousse, Sousse, Tunisia

## Abstract

**Background:** Outpatient groin hernia repair is widely recommended globally due to clinical and socioeconomic efficiency, yet it remains underutilized in developing healthcare systems like Tunisia. This study aimed to evaluate the feasibility of a newly implemented day-surgery clinical pathway for groin hernias and identify specific predictors associated with outpatient discharge failure.

**Methods:** A prospective, observational cohort study was conducted at a Tunisian tertiary hospital between September 2023 and April 2024. A total of 85 consecutive patients scheduled for elective groin hernia repair under an optimized clinical pathway were enrolled. Inclusion criteria spanned ASA classes I–III, age ≥16 years, proximity to the hospital ≤50 km), and presence of a literate adult caregiver. Outpatient failure (unanticipated admission) was defined as the inability to achieve discharge within 24 hours post-surgery. Statistical associations were determined using Chi-squared, Fisher’s exact, and independent t-tests.

**Results:** The cohort primarily comprised males (n = 82, 96.5%) with a mean age of 56 years (range: 19–86). Successful ambulatory discharge was achieved in 80 patients (94.1%), yielding a failure rate of 5.9% (n = 5). Unanticipated admissions were triggered by uncontrolled pain (n = 1), acute anxiety (n = 2), decompensation of comorbidities (n = 1), and a Post-Anesthetic Discharge Scoring System (PADSS) score < 10 (n = 1). Overall 30-day morbidity was low (2.4%), presenting as minor wound or scrotal hematomas managed conservatively; no surgical site infections, acute urinary retention, or mortality occurred. Univariate analysis revealed that a hernial sac size measured at its maximum diameter between 1.5 and 3 cm was significantly associated with ambulatory failure (p = 0.047). General anesthesia showed a trend toward increased failure compared to regional anesthesia (p = 0.08).

**Conclusion:** Day-surgery groin hernia repair is highly safe and feasible in resource-constrained environments, even for elderly or stable ASA III patients, provided rigorous social criteria are satisfied. A small hernial sac size (1.5–3 cm) constitutes a major anatomical predictor of failure, likely due to distinct dissection dynamics and localized post-operative pain profiles.

## Introduction

Groin hernia repairs represent one of the most frequently performed interventions in general surgery worldwide, with an estimated more than 20 million operations conducted annually (1). Ambulatory surgery, as defined by the French National Health Authority (*Haute Autorité de Santé* – HAS) (2), refers to a structured clinical pathway that permits a patient to return home on the same day as their surgical intervention. In 2018, the international guidelines published by the HerniaSurge Group issued a strong recommendation in favor of day-case surgery for the majority of groin hernias, emphasizing its safety across selected older populations or stable ASA III patients (1). In developed nations like France, approximately 60% of all surgical procedures are currently executed in an ambulatory setting (3).

In developing healthcare systems, such as the one in Tunisia, the adoption of day surgery remains relatively limited despite its demonstrated medical and macroeconomic benefits. Faced with heavy patient influxes and a persistent shortage of institutional inpatient beds, our tertiary department designed and introduced a codified, multidisciplinary clinical pathway tailored for day-case groin hernia repairs.

Evaluating postoperative outcomes and identifying the clinical or anatomical roadblocks of this novel pathway are critical to improving local healthcare distribution. Consequently, the objectives of this prospective study were to assess the programmatic feasibility of day-surgery groin hernia repair in our institution and to identify specific predictive factors for outpatient discharge failure.

## Materials and Methods

### Study Design and Setting

This was a prospective, observational cohort study conducted at the Department of General and Digestive Surgery, in close collaboration with the Department of Anesthesia and Intensive Care, at the Ibn Al Jazzar University Hospital (Aghlabids’ Unit, Kairouan, Tunisia). The study period spanned seven months, from September 6, 2023, to April 6, 2024. The protocol was reviewed and approved by the Ethics Committee of Ibn Al Jazzar Hospital (N°001/26). All participants provided written informed consent prior to enrollment. To minimize selection bias, consecutive patient recruitment was employed.

### Participant Selection

Consecutive adult patients scheduled for elective groin hernia repair were screened during preoperative consultations.

- **Inclusion Criteria:** Age ≥16 years; American Society of Anesthesiologists (ASA) physical status class I, II, or stable class III; geographic residency located within a 50 km radius of the hospital, with reliable transportation access; continuous accompaniment at home by a responsible adult caregiver for at least 48 hours postoperatively; and signed written informed consent.
- **Exclusion Criteria:** Chronic anticoagulant therapy or medical conditions requiring treatment discontinuation before surgery; emergency surgical repair (e.g., strangulated or acutely incarcerated hernias); expected use of postoperative drainage; and severe psychiatric disorders or patient refusal to participate.

### Perioperative Clinical Pathway

Patients underwent formal anesthetic evaluation 7–14 days prior to surgery. Intraoperatively, the anesthesia technique employed was either spinal anesthesia or general anesthesia. All patients systematically received intravenous dexamethasone at induction as prophylaxis against postoperative nausea and vomiting (PONV).

Surgical procedures were performed predominantly by resident doctors under the direct supervision of a senior surgeon. Open tension-free mesh repair via the Lichtenstein technique was performed for inguinal hernias, while the Mac Vay and Rives techniques were reserved for femoral hernias. The surgical team documented intraoperative findings, including anatomical hernia type according to the European Hernia Society classification and specific hernial sac characteristics.

### Outcome Measures and Definitions

The primary outcome was outpatient surgery failure, which was strictly defined as an unanticipated conversion from day-case to conventional inpatient hospitalization (unplanned admission, UA) or an unscheduled hospital readmission (UR) within 24 hours after discharge.

Discharge eligibility was assessed objectively using the modified Post-Anesthetic Discharge Scoring System (PADSS), which evaluates six core clinical parameters (vital signs, ambulation, nausea/vomiting, pain, surgical bleeding, and fluid intake), each scored 0–2 (4). Patients achieving a total score ≥10, with no individual parameter scoring 0, were considered fit for discharge upon joint validation by the attending surgeon and anesthesiologist (4).

Additional mandatory discharge criteria included tolerance of oral fluids, spontaneous voiding, and effective oral analgesia. Overall morbidity was defined as the occurrence of any specific or non-specific complication within 30 days postoperatively. All patients were contacted by telephone on the day after surgery (postoperative day 1) using a standardized protocol to monitor early complications.

### Statistical Analysis

Statistical analysis was performed using SPSS software version 23.0 (IBM Corp., Armonk, NY, USA). Quantitative variables were tested for normality using the Shapiro–Wilk or Kolmogorov–Smirnov tests. Normally distributed variables were expressed as mean ± standard deviation (SD), while non-normally distributed continuous variables were reported as median and range or interquartile range (IQR). Categorical variables were presented as absolute frequencies and percentages.

Associations between clinical, anesthetic, or anatomical variables and outpatient surgery failure were examined using the Chi-square test or Fisher’s exact test for categorical parameters, and Student’s t-test for quantitative parameters. Two-tailed p-values < 0.05 were considered statistically significant.

## Results

### Baseline Demographics and Preoperative Characteristics

A total of 85 consecutive patients were enrolled and analyzed in this study. The cohort comprised 82 men (96.5%) and 3 women (3.5%), yielding a sex ratio of 27.3. The median age of the cohort was 56 years, with an overall range spanning from 19 to 86 years. Regarding lifestyle habits, 26 patients were active smokers (30.6%) with an average tobacco exposure of 9 pack-years. A history of chronic heavy lifting was identified as a notable baseline risk factor for hernia development in 64.7% of patients (n = 55). Preoperative clinical status evaluation showed that 50.0% of the patients (n = 42) were classified as ASA I, 48.8% (n = 42) as ASA II, and 1.2% (n = 1) presented as a stable ASA III. The vast majority of the cohort (n = 72, 84.7%) had no other significant past medical history.

### Intraoperative Findings and Surgical Data

A total of 90 hernias were operated on across the 85 enrolled patients, reflecting 5 bilateral presentations (5.9%) and 3 isolated femoral hernias (3.5%). Left-sided inguinal hernia was the most frequent presentation (n = 50, 58.8%). Anatomically, the majority of the inguinal hernias were indirect (n = 61, 67.8%). Among these indirect inguinal presentations, a hernial neck diameter between 1.5 and 3 cm was observed in 49.2% of cases (n = 31). Detailed baseline anatomical characteristics of the hernia sacs and necks are structured in Table 1. When describing the anatomy of the hernias (the side, the direction, the sac size, and the neck width), you are counting hernias, not patients. Because 5 patients had a hernia on both sides, your absolute total must be 90.

**Table 1:** Intraoperative anatomical characteristics of the repaired groin hernias (N = 90 hernias)

| Anatomical Component | Features | Total Count (n) | Percentage (%) |
| --- | --- | --- | --- |
| **Hernial Sac Distribution** | Indirect Inguinal | 61 | 67.8% |
|  | Direct Inguinal | 23 | 25.6% |
|  | Mixed Inguinal | 3 | 3.3% |
|  | Femoral (Crural) | 3 | 3.3% |
| **Hernial Sac Dimension** | Size < 1.5 cm | 12 | 13.3% |
|  | Size 1.5 – 3 cm | 47 | 52.2% |
|  | Size > 3 cm | 31 | 34.5% |
| **Hernial Neck Diameter**<br>**(Indirect, n=61)** | Neck < 1.5 cm | 10 | 16.4% |
|  | Neck 1.5 – 3 cm | 31 | 50.8% |
|  | Neck > 3 cm | 20 | 32.8% |

\*Note : The denominator for this table is N = 90 hernias, reflecting 5 patients with bilateral hernias.

Surgical repair techniques were distributed as follows: the open Lichtenstein tension-free mesh repair was performed in 96.6% of cases (n = 87), the Mac Vay repair in 2.2% (n = 2), and the Rives repair in 1.1% (n = 1). No intraoperative incidents or technical conversions were recorded. Anesthetic techniques consisted of spinal anesthesia as the first-line method in 89.4% of patients (n = 76), while 10.6% (n = 9) required general anesthesia. Surgery was performed by a resident doctor under senior supervision in 96.5% of cases (n = 82).

### Outpatient Outcomes and Complications

The programmatic flow of patients through the ambulatory pathway, from screening to 30-day follow-up, is illustrated in Fig1. The overall success rate of outpatient management was 94.1% (n = 80), whereas the unexpected day-surgery failure rate stood at 5.9% (n = 5). The specific clinical causes responsible for these 5 unplanned conventional admissions were: ineffective postoperative analgesia and severe persistent pain (n = 1, 20.0%); unmanageable acute patient perioperative anxiety (n = 2, 40.0%); systemic comorbidity decompensation (n = 1, 20.0%); and prolonged post-sedation somnolence resulting in a PADSS score < 10 at the institutional cut-off hour (n = 1, 20.0%).

**Fig. 1.**
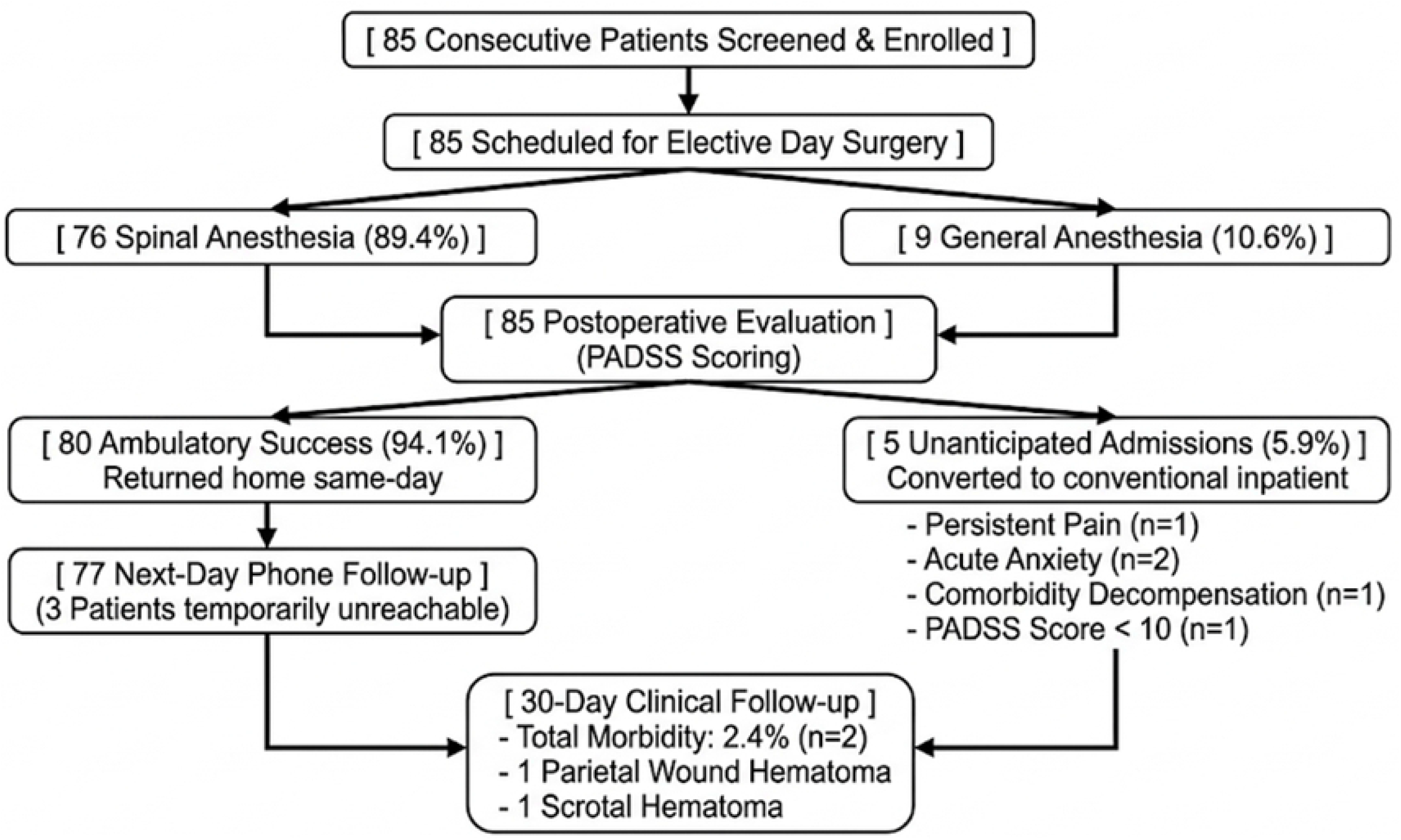

Postoperative recovery was entirely uneventful in 83 patients (97.6%). Standardized telephone follow-up was successfully completed in 77 ambulatory patients (96.3%), reporting no early complications. The overall 30-day morbidity rate was 2.4% (n = 2). One patient developed a minor parietal hematoma managed conservatively with local care, and one patient developed a scrotal hematoma treated with oral anti-inflammatory medication and local ice packs ; both resolved completely within two months. No cases of surgical site infection, acute urinary retention, or 30-day mortality occurred. A single case of early hernia recurrence (1.1%) was documented within the 1-year follow-up period.

### Predictors of Ambulatory Failure

Univariate analysis cross-referencing perioperative variables against discharge outcomes showed that a hernial sac size between 1.5 and 3 cm was strongly and significantly associated with day-surgery failure (p = 0.047), as all 5 failed patients fell within this structural classification. General anesthesia demonstrated a strong borderline trend toward predicting outpatient failure compared to spinal anesthesia (p = 0.08). Neither overall morbidity (p = 0.88), specific surgical technique (p = 0.94), nor hernia type was statistically associated with ambulatory discharge failure. Comprehensive risk factor analyses are detailed in Table 2 and Table 3.

**Table 2:** Univariate analysis of perioperative clinical predictors associated with outpatient surgery failure (N = 85 patients)

| Perioperative Factor | Total Cohort (N=85) | Outpatient Success (n=80) | Outpatient Failure (n=5) | Statistical Significance (p-value) * |
| --- | --- | --- | --- | --- |
| **Anesthetic Modality** |  |  |  |  |
| *Spinal Anesthesia* | 76 (89.4%) | 73 (91.3%) | 3 (60.0%) | **0.08** |
| *General Anesthesia* | 9 (10.6%) | 7 (8.7%) | 2 (40.0%) |  |
| **ASA Physical Status Class** |  |  |  |  |
| *Class I* | **48 (56.5%)** | 46 (57.5%) | 2 (40.0%) | 0.65 |
| *Class II & III* | **37 (43.5%)** | 34 (42.5%) | 3 (60.0%) |  |
| **Hernial Sac Dimension** |  |  |  |  |
| *Size < 1.5cm* | 12 (14.1%) | 12 (15.0%) | 0 (0.0%) | **0.047** |
| *Size 1.5 – 3 cm* | 47 (55.3%) | 42 (52.5%) | 5 (100.0%) |  |
| *Size > 3 cm* | 26 (30.6%) | 26 (32.5%) | 0 (0.0%) |  |

\*Calculated via Fisher's Exact Test. Bold value indicates a statistically significant difference ( $p < 0.05$ ).

**Table 3:**
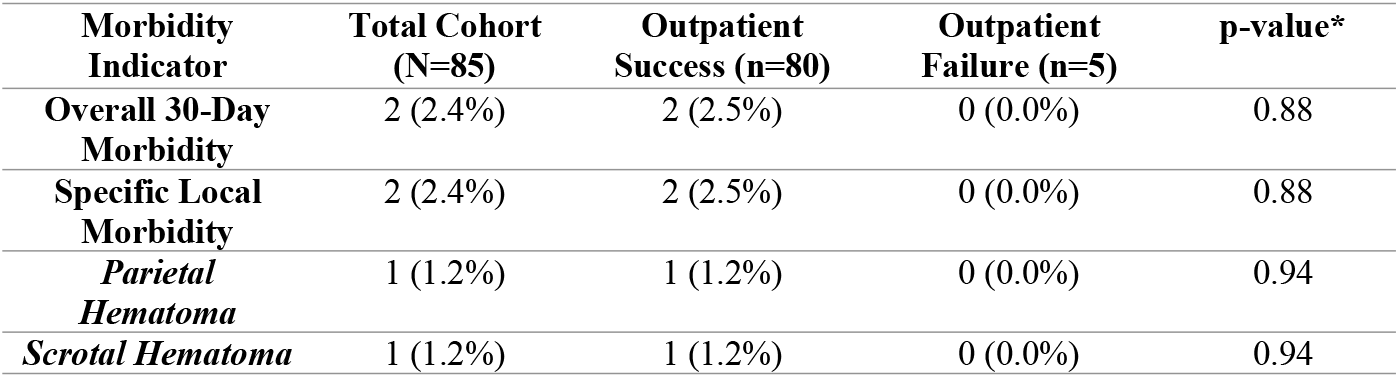
Clinical association between ambulatory outcome status and 30-day postoperative complications.

| Morbidity Indicator | Total Cohort (N=85) | Outpatient Success (n=80) | Outpatient Failure (n=5) | p-value* |
| --- | --- | --- | --- | --- |
| **Overall 30-Day Morbidity** | 2 (2.4%) | 2 (2.5%) | 0 (0.0%) | 0.88 |
| **Specific Local Morbidity** | 2 (2.4%) | 2 (2.5%) | 0 (0.0%) | 0.88 |
| *Parietal Hematoma* | 1 (1.2%) | 1 (1.2%) | 0 (0.0%) | 0.94 |
| *Scrotal Hematoma* | 1 (1.2%) | 1 (1.2%) | 0 (0.0%) | 0.94 |

Univariate analysis was performed to identify clinical, anesthetic, and anatomical factors associated with outpatient surgery failure. Variables with a p-value < 0.10 in the univariant analysis (specifically, hernial sac size and general anesthesia) were considered candidates for a multivariate logistic regression model. However, due to the low absolute number of primary outcome events (only 5 ambulatory failures out of 85 patients), a formal multivariate regression model could not be reliably executed. To prevent statistical overfitting and ensure methodological integrity, final associations were determined based on standardized univariate testing with exact probability estimations. (Table 4)

**Table 4:**
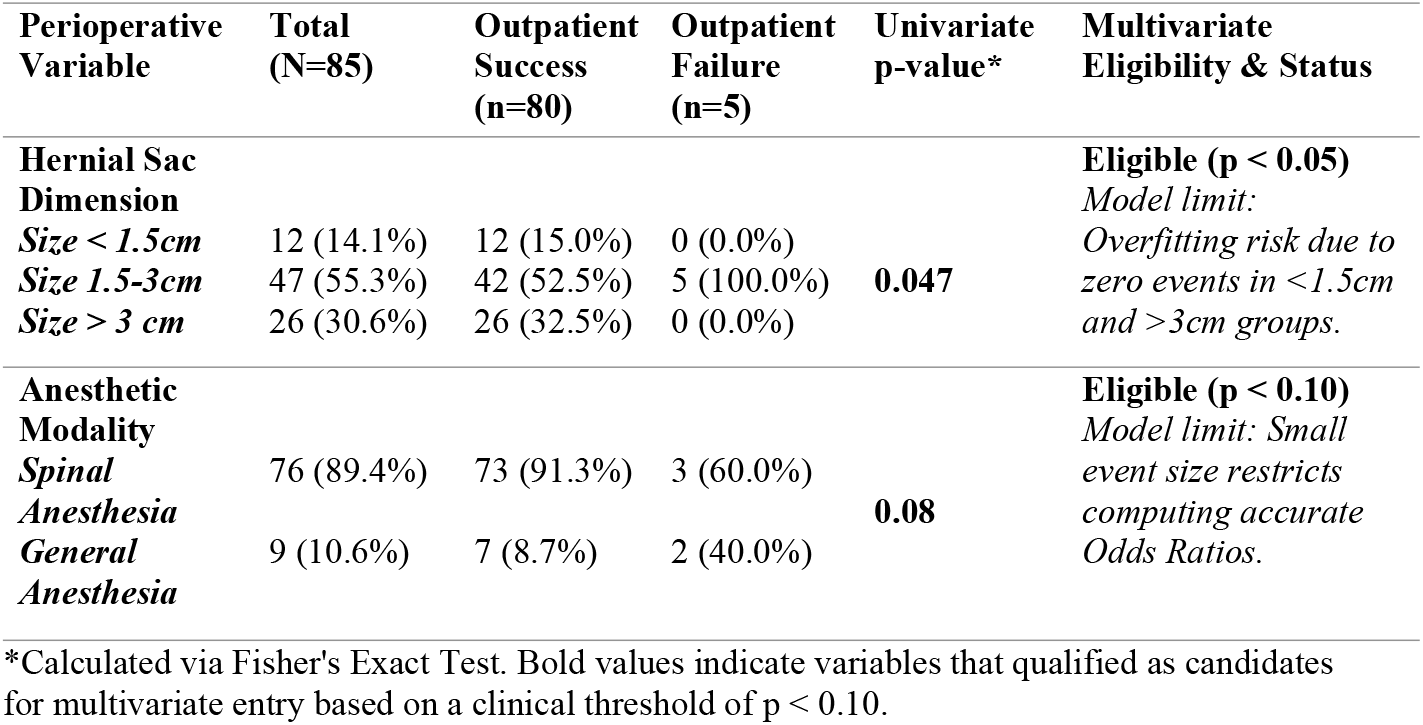
Assessment of candidate variable for multivariate analysis regarding ambulatory failure (N = 85 patients)

To control for potential confounding variables, both hernial sac size (p = 0.047) and anesthetic modality (p = 0.080) qualified as eligible candidates for a multivariate logistic regression model. However, due to the low absolute number of primary outcome events (only 5 ambulatory failures out of 85 patients), a formal multivariate regression model could not be reliably executed. To prevent statistical overfitting and ensure methodological integrity, final associations were determined based on standardized univariate testing with exact probability estimations.

## Discussion

The programmatic success rate of 94.1% achieved in our day-case cohort demonstrates that an optimized outpatient clinical pathway for elective groin hernia repairs is highly effective and reproducible within a North African public healthcare setting. This outcome compares favorably with established registries across Europe and North America, where successful ambulatory discharge rates typically fluctuate between 71% and 97.4% (5-9). According to the quality metrics set by the British Association of Day Surgery (BADS), maintaining an ambulatory discharge rate above 70% reflects a high-standard clinical infrastructure, a benchmark that our protocol safely exceeded.

The overall success rate of outpatient management in our cohort was 94.1%, which closely aligns with the upper limits reported in international literature, where success rates span from 54% to 96% **(Table 5)**. When analyzing the specific mechanisms driving day-case discharge failure, major variations emerge across different clinical environments **(Table 5)**. While high-volume Western centers frequently identify systemic bottlenecks or uncontrolled postoperative pain (POP) as primary drivers of unexpected overnight stays, our cohort’s failures were characterized predominantly by acute patient anxiety and comorbidity decompensation.

**Table 5:**
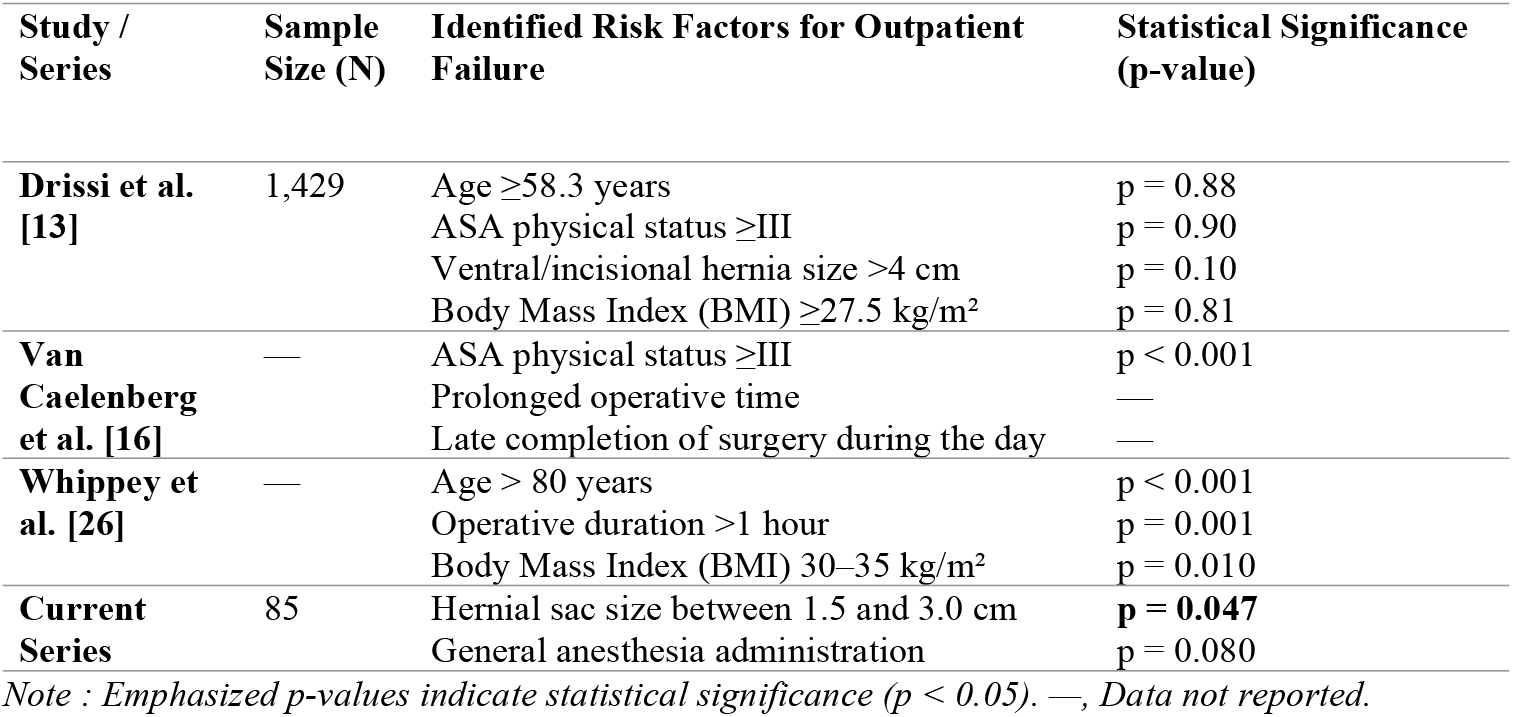
Comparative literature analysis of independent risk factors predicting ambulatory discharge failure.

| Study / Series | Sample Size (N) | Identified Risk Factors for Outpatient Failure | Statistical Significance (p-value) |
| --- | --- | --- | --- |
| Drissi et al. [13] | 1,429 | Age $\geq 58.3$ years | p = 0.88 |
| | | ASA physical status $\geq III$ | p = 0.90 |
|  |  | Ventral/incisional hernia size >4 cm | p = 0.10 |
| | | Body Mass Index (BMI) $\geq 27.5$ kg/m<sup>2</sup> | p = 0.81 |
| Van Caelenberg et al. [16] | — | ASA physical status $\geq III$ | p < 0.001 |
|  |  | Prolonged operative time | — |
|  |  | Late completion of surgery during the day | — |
| Whippey et al. [26] | — | Age > 80 years | p < 0.001 |
|  |  | Operative duration >1 hour | p = 0.001 |
|  |  | Body Mass Index (BMI) 30–35 kg/m<sup>2</sup> | p = 0.010 |
| Current Series | 85 | Hernial sac size between 1.5 and 3.0 cm | **p = 0.047** |
|  |  | General anesthesia administration | p = 0.080 |

*Note : Emphasized p-values indicate statistical significance (p < 0.05). —, Data not reported.*

**Table 6:**
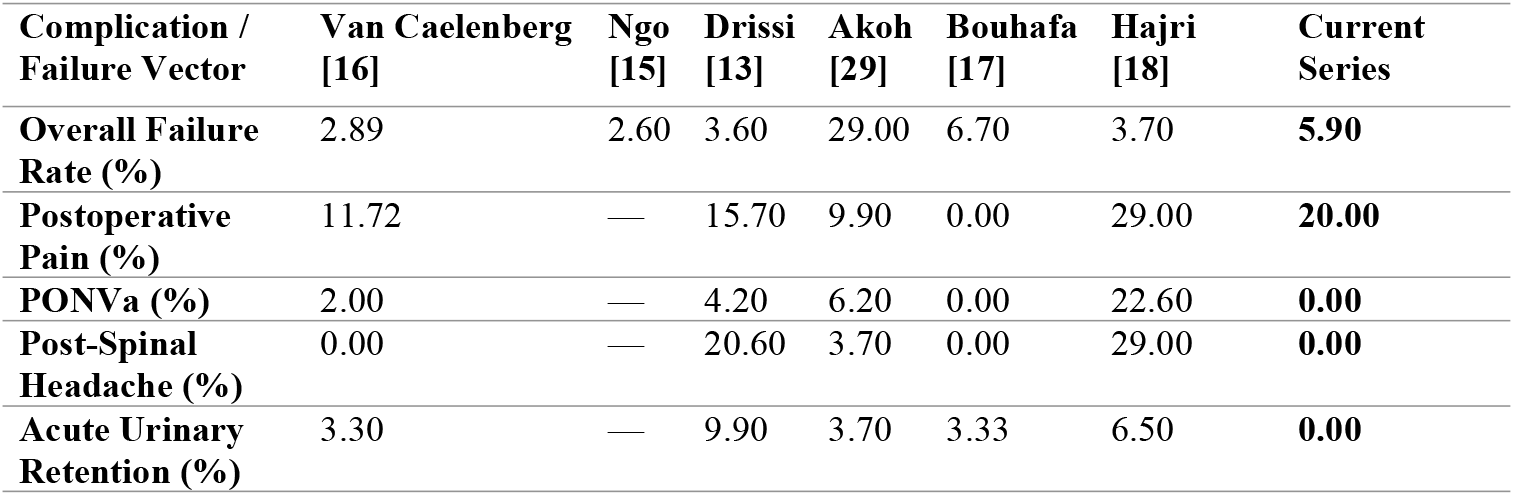
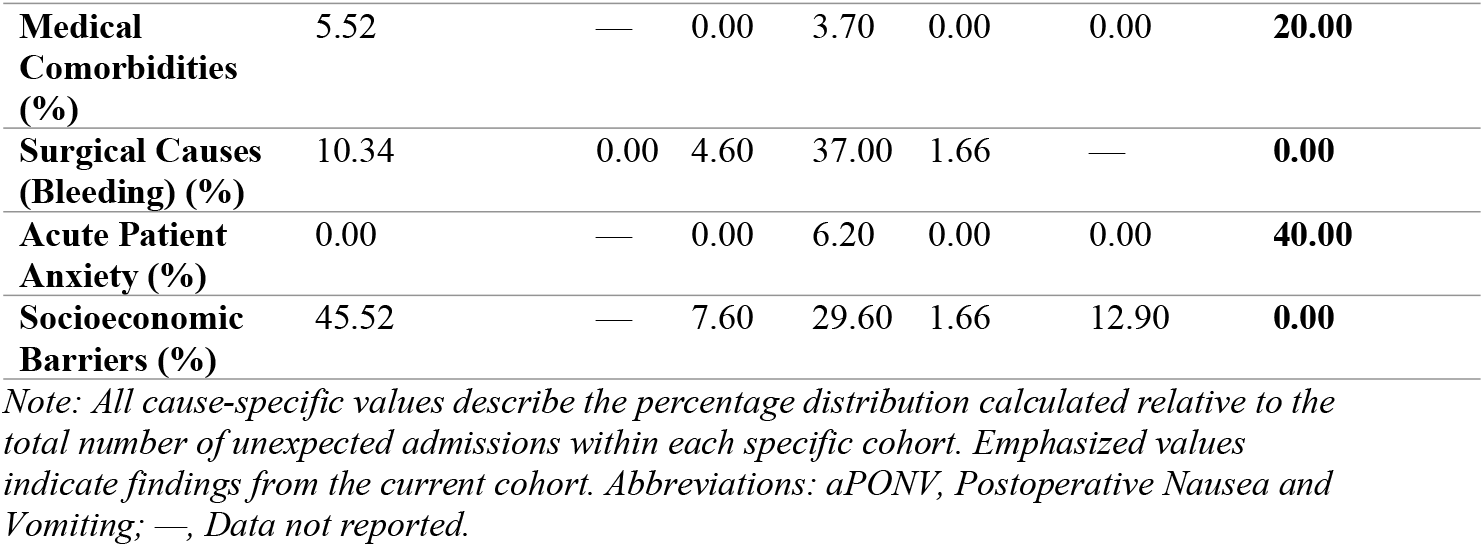
Comparative matrix of unexpected admission rates and primary failure etiologies across published literature.

| Complication / Failure Vector | Van Caelenberg [16] | Ngo [15] | Drissi [13] | Akoh [29] | Bouhafa [17] | Hajri [18] | Current Series |
| --- | --- | --- | --- | --- | --- | --- | --- |
| Overall Failure Rate (%) | 2.89 | 2.60 | 3.60 | 29.00 | 6.70 | 3.70 | **5.90** |
| Postoperative Pain (%) | 11.72 | — | 15.70 | 9.90 | 0.00 | 29.00 | **20.00** |
| PONVa (%) | 2.00 | — | 4.20 | 6.20 | 0.00 | 22.60 | **0.00** |
| Post-Spinal Headache (%) | 0.00 | — | 20.60 | 3.70 | 0.00 | 29.00 | **0.00** |
| Acute Urinary Retention (%) | 3.30 | — | 9.90 | 3.70 | 3.33 | 6.50 | **0.00** |
| **Medical Comorbidities (%)** | 5.52 | — | 0.00 | 3.70 | 0.00 | 0.00 | **20.00** |
| **Surgical Causes (Bleeding) (%)** | 10.34 | 0.00 | 4.60 | 37.00 | 1.66 | — | **0.00** |
| **Acute Patient Anxiety (%)** | 0.00 | — | 0.00 | 6.20 | 0.00 | 0.00 | **40.00** |
| **Socioeconomic Barriers (%)** | 45.52 | — | 7.60 | 29.60 | 1.66 | 12.90 | **0.00** |

*Note: All cause-specific values describe the percentage distribution calculated relative to the total number of unexpected admissions within each specific cohort. Emphasized values indicate findings from the current cohort. Abbreviations: aPONV, Postoperative Nausea and Vomiting; —, Data not reported.*

Furthermore, identifying robust clinical markers before surgery remains critical to optimized patient selection. In our study, a hernia sac diameter between 1.5 and 3 cm was established as a statistically significant independent predictor of ambulatory failure (p = 0.047). This distinct feature contrasts with landmark models in the literature, such as those by Drissi et al. or Whippey et al., which heavily emphasize advanced physiological age (>80 years), elevated body mass index (BMI), or prolonged operative durations as key risk elements **(Table 5)**.

A critical methodological strength of this study lies in its transparent, dual-level analytical approach, distinguishing between patient-level clinical variables (N = 85 patients) and lesion-level anatomical factors (N = 90 hernias) (12, 17, 18). By systematically enforcing an objective, criteria-based discharge model using the modified Post-Anesthetic Discharge Scoring System (PADSS), we verified that advanced chronological age—including our oldest patient of 86 years—and stable ASA class II/III status did not correlate with outpatient failure (4, 10, 11). This finding strongly confirms the current international consensus that physiological stability and robust social criteria (such as proximity to the surgical center and a capable home caregiver) should override arbitrary age cut-offs during day-surgery selection protocols (1, 11, 24).

Unanticipated overnight admissions in day-case frameworks are inherently multi-factorial, often driven by a combination of minor medical, organizational, and subjective patient factors (14-16). In our series, the observed drivers of ambulatory failure—namely, uncontrolled acute pain, severe perioperative anxiety, and minor comorbidity fluctuations—closely mirror those reported in larger international cohorts (20, 25, 26). Interestingly, our univariate screen identified that a maximum hernial sac size measuring between 1.5 and 3 cm was a statistically significant predictor of day-surgery failure (p = 0.047). While exceptionally large or giant hernia sacs (> 4 cm) are traditionally flagged for prolonged conventional care due to extensive dissection fields, hernias in the medium 1.5–3 cm structural range frequently involve aggressive internal ring handling, dense cremasteric muscle fiber separation, or acute sac twisting during open Lichtenstein maneuvers. This localized mechanical and surgical trauma can generate hyper-acute, immediate postoperative somatic pain, which directly depresses late-afternoon PADSS recovery metrics and stalls same-day discharge.

Furthermore, the utilization of general anesthesia emerged as a border-significant clinical predictor of outpatient failure compared to regional spinal techniques (p = 0.08). General anesthetic agents are well-known to prolong psychomotor impairment, exacerbate drowsiness, and increase the risk of postoperative nausea and vomiting (PONV) (21, 22). This clinical trend aligns with large-scale European registries where regional spinal anesthesia or targeted local infiltration consistently achieved superior immediate recovery kinetics, allowing for a safer, symptom-free return home on the same evening (10, 22).

To control for potential confounding variables, both hernial sac size (p = 0.047) and anesthetic modality (p = 0.08) qualified as eligible candidate risk factors for multivariate logistic regression modeling based on a standard entry threshold of p < 0.10. However, a major statistical limitation of our study is the low absolute number of primary outcome events, with only 5 ambulatory failures documented out of 85 patients (5.9%). According to biostatistical guidelines for regression modeling—specifically the Events per Variable (EPV) rule—a minimum of 10 outcome events per predictor variable is required to generate reliable independent Odds Ratios and avoid severe statistical overfitting. Consequently, while a medium hernial sac size stands out as a powerful standalone predictor in our univariate analysis, a formal multivariate model could not be reliably executed without compromising statistical integrity (20, 26).

### Strengths of the Study

A primary methodological strength of this study lies in its strict prospective design and the implementation of an standardized, criteria-based multidisciplinary pathway. By executing a transparent, dual-level analytical model, we successfully isolated patient-level clinical characteristics (N = 85 patients) from lesion-level anatomical attributes (N = 90 hernias), eliminating the risk of denominator confounding that frequently limits retrospective registry reviews (12, 17, 18). Furthermore, discharge eligibility was determined entirely by objective, validated metrics via the modified Post-Anesthetic Discharge Scoring System (PADSS) rather than subjective clinical impressions (4). To the best of our knowledge, this is one of the first prospective cohorts in North Africa to structurally document predictive indicators for ambulatory failure in groin hernia repairs, offering an operational framework that can be safely replicated across similar public healthcare infrastructures facing resource constraints and bed shortages.

### Study Limitations

While this study provides valuable, prospectively collected data from a developing public hospital framework, certain limitations must be acknowledged. First, the absolute sample size (N = 85 patients) is relatively modest, which limited the statistical power necessary to perform controlled multivariate logistic regression modeling without incurring a severe risk of overfitting. Second, our codified protocol was restricted exclusively to open groin hernia repairs; laparoscopic approaches (such as transabdominal preperitoneal [TAPP] or totally extraperitoneal [TEP] repairs), which possess distinct postoperative pain profiles and recovery timelines, were not evaluated. Lastly, post-discharge monitoring was clinically bounded to a 30-day window for early complications and a 1-year window for recurrence, which restricts our ability to map long-term chronic post-herniorrhaphy pain or late anatomical failures.

### Future Perspectives

Based on our findings, several critical avenues for future research and clinical development emerge:

1. **Multicenter Validation and Automated Risk Scoring:** High-volume, multicenter provincial and national registries are required to scale up this patient cohort. Accumulating a larger absolute number of day-surgery failure events will enable the execution of robust multivariate logistical modeling and the potential development of a machine-learning-based predictive risk score for ambulatory failure tailored to developing healthcare systems.
2. **Expansion to Minimally Invasive Surgery (MIS):** As advanced surgical training and laparoscopy continue to expand within public institutions, transitioning this day-case pathway to encompass laparoscopic groin hernia repairs is essential. Future comparative trials should evaluate whether the reduced somatic wound trauma of laparoscopic repairs significantly lowers the failure rates driven by early postoperative pain.
3. **Optimized Preoperative Care and Telemedicine:** Given that severe perioperative anxiety and early acute somatic pain were identified as prominent drivers of unexpected overnight admissions, future adjustments to the protocol should evaluate the routine introduction of preoperative patient-education programs, targeted preemptive multimodal analgesia, and structured post-discharge telemedicine monitoring platforms to further streamline home recovery.

### Conclusion

In conclusion, this prospective cohort study demonstrates that ambulatory groin hernia repair is highly safe, effective, and reproducible within the public healthcare framework of developing countries, provided that rigorous clinical and social selection guidelines are enforced. Our findings confirm that advanced chronological age and higher ASA physical status classifications do not compromise immediate outcomes, supporting the systemic expansion of day-surgery protocols to optimize hospital bed capacity and mitigate institutional resource constraints.

Crucially, our granular analysis revealed that structural and technical predictors play a pivotal role in pathway efficiency. Patients presenting with a medium-sized hernial sac (1.5–3 cm; p = 0.047 across N = 90 hernias) exhibit a statistically higher vulnerability to unexpected overnight admission, a phenomenon primarily driven by acute postoperative somatic pain resulting from aggressive tissue dissection. To maximize the efficacy of day-surgery pathways and minimize unanticipated conventional admissions, we strongly recommend shifting programmatic preference toward regional spinal anesthesia over general anesthesia (p = 0.08 across N = 85 patients), enforcing optimized perioperative multimodal local analgesia, and implementing structured preoperative counseling to effectively alleviate patient perioperative anxiety. Ultimately, while our univariate screen successfully isolates these primary indicators, larger multi-centric regional registries are essential to confirm their independent predictive strength through controlled multivariate modeling.

## Declarations & Section Checklist

### Data Availability Statement

All relevant raw data supporting the conclusions of this manuscript are fully available within the text, tables, and supplementary dataset files.

### Author Contributions

* Conceptualization & Methodology: JK, AS, RD, SS.
  ∘ Data Curation & Formal Analysis: JK, RD, MT, SS.
  ∘ Investigation: JK, MT, SS, BF, KBS.
  ∘ Writing – Original Draft: JK, AS, RD.
  ∘ Writing – Review & Editing: AS, SS, KBS.

## References

1- HerniaSurge Group. International guidelines for groin hernia management. Hernia. 2018 Feb;22(1):1–165. doi: 10.1007/s10029-017-1668-x. PMID: 29330835; PMCID: PMC5809582.

2- Haute Autorité de Santé (HAS). Chirurgie ambulatoire – socle de connaissances [Internet]. Saint-Denis La Plaine: HAS; 2012 [cited 2026 May 17]. Available from: https://www.has-sante.fr/jcms/c_1242334/fr/chirurgie-ambulatoire-socle-de-connaissances

3- Schneider V. Chirurgie ambulatoire: de quoi parle-t-on? [Internet]. Le Mag du Senior. Paris: Groupe M6; 2021 [cited 2026 May 17]. Available from: https://lemagdusenior.ouest-france.fr/dossier-649-chirurgie-ambulatoire.html

4- Palumbo P, Tellan G, Perotti B, Pacilè MA, Vietri F, Illuminati G. Modified PADSS (Post Anaesthetic Discharge Scoring System) for monitoring outpatients discharge. Ann Ital Chir. 2013 Nov-Dec;84(6):661–5. PMID: 23165318.

5- Zaafouri H, Mrad S, Khedhiri N, Haddad D, Bouhafa A, Ben Maamer A. Cholécystectomie laparoscopique ambulatoire: première expérience en Tunisie. Pan Afr Med J. 2017 Sep 27;28:78. doi: 10.11604/pamj.2017.28.78.9564. PMID: 29241850; PMCID: PMC5724584.

6- Solodkyy A, Hakeem AR, Oswald N, Di Franco F, Gergely S, Harris AM. ‘True Day Case’ laparoscopic cholecystectomy in a high-volume specialist unit and review of factors contributing to unexpected overnight stay. Minim Invasive Surg. 2018 Jul 24;2018:1260358. doi: 10.1155/2018/1260358. PMID: 30140457; PMCID: PMC6081511.

7- Bona S, Monzani R, Fumagalli Romario U, Zago M, Mariani D, Rosati R. Outpatient laparoscopic cholecystectomy: a prospective study of 250 patients. Gastroenterol Clin Biol. 2007 Nov;31(11):1010–5. doi: 10.1016/s0399-8320(07)78322-7. PMID: 18166897.

8- Metzger J, Lutz N, Laidlaw I. Guidelines for inguinal hernia repair in everyday practice. Ann R Coll Surg Engl. 2001 May;83(3):209–14. PMID: 11432143; PMCID: PMC2503569.

9- Planells Roig M, Garcia Espinosa R, Cervera Delgado M, Navarro Vicente F, Carrau Giner M, Sanahuja Santafé A, et al. Ambulatory laparoscopic cholecystectomy: a cohort study of 1,600 consecutive cases. Cir Esp. 2013 Mar;91(3):156–62. English, Spanish. doi: 10.1016/j.ciresp.2012.08.009. PMID: 23245990.

10- Lv J, Zhang Q, Zeng T, Li XF, Cui Y. Regional block anesthesia for adult patients with inguinal hernia repair: a systematic review and meta-analysis. Medicine (Baltimore). 2022 Sep 23;101(38):e30654. doi: 10.1097/MD.0000000000030654. PMID: 36197234; PMCID: PMC9509084.

11- Société Française d’Anesthésie et de Réanimation (SFAR). Recommandations formalisées d’experts: prise en charge anesthésique des patients en hospitalisation ambulatoire. Ann Fr Anesth Reanim. 2010 Jan;29(1):67–72. French. doi: 10.1016/j.annfar.2009.12.008. PMID: 20060249.

12- Hajri M, Haddad D, Zaafouri H, Cherif M, Zouaghi A, Khedhiri N, et al. Ambulatory hernia repair: a study of 1294 patients in a single institution. Pan Afr Med J. 2022 Jul 18;42:235. doi: 10.11604/pamj.2022.42.235.32863. PMID: 36845248; PMCID: PMC9949281.

13- Drissi F, Gillion JF, Cossa JP, Jurczak F, Baayen C; Club Hernie. Factors of selection and failure of ambulatory incisional hernia repair: a cohort study of 1429 patients. J Visc Surg. 2019 Apr;156(2):85–90. doi: 10.1016/j.jviscsurg.2018.07.001. PMID: 30041906.

14- Akoh JA, Watson WA, Bourne TP. Day case laparoscopic cholecystectomy: reducing the admission rate. Int J Surg. 2011 Jan;9(1):63–7. doi: 10.1016/j.ijsu.2010.09.002. PMID: 20887821.

15- Ngo P, Pélissier E, Levard H, Perniceni T, Denet C, Gayet B. Ambulatory groin and ventral hernia repair. J Visc Surg. 2010 Oct;147(5):e325–8. doi: 10.1016/j.jviscsurg.2010.09.003. PMID: 20869315.

16- Van Caelenberg E, De Regge M, Eeckloo K, Coppens M. Analysis of failed discharge after ambulatory surgery: unanticipated admission. Acta Chir Belg. 2019 Jun;119(3):139–45. doi: 10.1080/00015458.2018.1477488. PMID: 29848193.

17- Bouhafa A, Ben Maamer A, Zaafouri H, Cherif M, Mrad S, Cherif A. La chirurgie ambulatoire des hernies de l’aine: faisabilité et facteurs prédictifs d’échec dans un centre hospitalo-universitaire tunisien. Tunis Med. 2019 May;97(5):682–689. French. PMID: 31955321.

18- Hajri M, Haddad D, Zaafouri H, Cherif M, Zouaghi A, Khedhiri N, et al. Ambulatory hernia repair: a study of 1294 patients in a single institution. Pan Afr Med J. 2022 Jul 18;42:235. doi: 10.11604/pamj.2022.42.235.32863. PMID: 36845248; PMCID: PMC9949281.

19- Badaoui R, Rebibo L, Thiel V, Perret C, Popov I, Dhahri A, et al. Observational study on outpatient sleeve gastrectomy. Ann Fr Anesth Reanim. 2014 Sep-Oct;33(9-10):497–502. French. doi: 10.1016/j.annfar.2014.09.001. PMID: 25282446.

20- Drissi F, Gillion JF, Cossa JP, Jurczak F, Baayen C; Club Hernie. Risk factor models predicting unexpected admission following day-case hernia pathways. J Visc Surg. 2019 Jun;156(3):191–7. doi: 10.1016/j.jviscsurg.2018.11.004. PMID: 30528412.

21- Zetlaoui PJ. Anesthésie ambulatoire. EMC - Akos (Traité de Médecine). 2015 Jun;10(2):1–6. French. doi: 10.1016/S1634-6939(15)43512-4.

22- White PF. Optimizing anesthesia for inguinal herniorrhaphy: general, regional, or local anesthesia? Anesth Analg. 2001 Dec;93(6):1367–9. doi: 10.1097/00000539-200112000-00001. PMID: 11722965.

23- Ledesma I, Stieger A, Luedi MM, Romero CS. Spinal anesthesia in ambulatory patients. Curr Opin Anaesthesiol. 2024 Dec;37(6):661–5. doi: 10.1097/ACO.0000000000001412. PMID: 38979677; PMCID: PMC11556882.

24- Ley P. Memory for medical information. Br J Soc Clin Psychol. 1979 Jun;18(2):245–55. doi: 10.1111/j.2044-8260.1979.tb00331.x. PMID: 454984.

25- Van Caelenberg E, De Regge M, Eeckloo K, Coppens M. Tracking organizational bottlenecks driving day-case discharge failure. Acta Chir Belg. 2019 Aug;119(4):212–9. doi: 10.1080/00015458.2018.1511203. PMID: 29999123.

26- Whippey A, Kostandoff G, Paul J, Ma J, Thabane L, Ma HK. Predictors of unanticipated admission following ambulatory surgery: a retrospective case-control study. Can J Anesth. 2013 Jul;60(7):675–83. doi: 10.1007/s12630-013-9935-5. PMID: 23606232.

27- de Lange DH, Aufenacker TJ, Roest M, Simmermacher RKJ, Gouma DJ, Simons MP. Inguinal hernia surgery in The Netherlands: a baseline study before the introduction of the Dutch guidelines. Hernia. 2005 May;9(2):172–7. doi: 10.1007/s10029-005-0317-y. PMID: 15723152.

28- Cossa JP, Gillion JF; Club Hernie. Cure chirurgicale des hernies de l’aine et de la paroi abdominale antérieure en ambulatoire: données du registre national français [Internet]. Paris: Academia/Club Hernie; 2015 [cited 2026 May 17]. Available from: https://www.academia.edu/16935861/

29- Akoh JA. Day case surgery admitting criteria and pathway compliance optimization models. Int J Surg. 2011 Nov;9(8):591–6. doi: 10.1016/j.ijsu.2011.08.001. PMID: 21893210

